# Determinants and experiences of care-seeking for childhood pneumonia in a rural Indian setting: A mixed-methods study

**DOI:** 10.1101/2025.04.04.25325289

**Authors:** Barsha Gadapani Pathak, Sarmila Mazumder, Yasir Bin Nisar, Aditya Bhatt, Mandeep Singh, Tarun Madhur, Ingvild Fossgard Sandøy

## Abstract

**Introduction:** Pneumonia is a leading cause of under-five mortality, with 30 million annual cases in India. Despite national guidelines, significant barriers to timely and appropriate care-seeking persist. Caregivers often face financial constraints, lack of awareness, mistrust in government facilities, and a preference for non-registered medical practitioners (non-RMP), delaying effective treatment. This study explores care-seeking behaviors, associated sociodemographic factors, and barriers to access to appropriate healthcare for childhood pneumonia in rural India.

**Methods:** This study is part of a broader implementation research (IR) initiative and represents its formative phase. This mixed-methods study was conducted in Palwal district, Haryana, covering 42 villages (population: 107,440). A cross-sectional survey identified suspected pneumonia cases among 9,593 under-five children through house-to-house visits using a structured checklist. Data on sociodemographic characteristics, health insurance, care-seeking patterns, and provider preferences were collected. Directed Acyclic Graphs (DAGs) identified potential confounders in the association between care-seeking behaviour and key exposure variables. Additionally, qualitative in-depth interviews explored caregivers’ perceptions, decision-making, and healthcare barriers to pneumonia management. Quantitative data were analysed using logistic regression, while qualitative data were thematically analyzed using the Three Delays Model. Suspected under-five pneumonia cases’ caregivers and families were actively engaged in this formative phase to inform Phase II implementation strategies of broader IR, ensuring community-driven and contextually relevant strategies.

**Results:** Among 231 suspected pneumonia cases, 97% of caregivers sought medical care, but 71% consulted non-RMPs, and only 3.6% visited government facilities. Seeking appropriate care was associated with higher maternal education (AOR 6.5, 95% CI: 2.7, 9.7) and higher wealth index (AOR 1.7, 95% CI: 1.0, 2.6). Thematic analysis revealed delays due to symptom misinterpretation, reliance on home remedies, financial constraints, gender biases, mistrust in public healthcare services, and logistical barriers.

**Conclusion:** Despite high care-seeking rates, provider preferences, socio-cultural factors, and systemic barriers delay appropriate pneumonia treatment. Addressing these challenges requires improving awareness, enhancing public healthcare trust, and strengthening frontline health worker engagement. This study underscores the role of structured beneficiary involvement in refining pneumonia management strategies to ensure sustainable, community-driven interventions.

## PATIENT OR PUBLIC CONTRIBUTION

This study is part of an ongoing implementation research (IR) aimed at improving the effective coverage of childhood pneumonia management in a low-resource setting. Structured engagement with primary caregivers of under-five children, mothers, fathers, family members, community members, and local community stakeholders/representatives e.g. local leaders, village heads etc. has been integrated at multiple stages to ensure the relevance and applicability of its findings.

The current study is part of Phase I (formative research) of the IR, where primary caregivers and family members participated in a needs assessment, providing critical insights into the barriers and facilitators influencing care-seeking for childhood pneumonia in a rural low-to-middle socioeconomic setting. Their inputs have informed the refinement of study tools and the development of mitigation strategies for the logic and implementation model. As the research progresses into Phase II (model development and implementation), the community continues to play an integral role in providing feedback on the feasibility and appropriateness of proposed strategies. This ongoing feedback loop is assessing how effectively these strategies strengthen linkages between the healthcare system, and the community, foster an active local needs assessment mechanism among healthcare providers, and enhance demand generation for appropriate pneumonia care-seeking. These iterative refinements ensure that the implementation strategies remain responsive to the evolving needs of the community. In the forthcoming Phase III, which will focus on scaling up the finalized implementation model, strategies will be adapted to further improve care-seeking for under-five children. Continuous engagement with caregivers and local community representatives, including Panchayati Raj Institution (PRI) members, will be central to refining these strategies. Additionally, during the dissemination phase, key findings will be shared with caregivers, community members, and PRI representatives, facilitating discussions on study implications and informing future policy and programmatic decisions. Their ongoing involvement will help contextualize findings and enhance the long-term sustainability of strategies aimed at improving pneumonia care-seeking behaviours and effective management in rural India.

## INTRODUCTION

Pneumonia is the leading infectious cause of death among children under five worldwide, claiming over 800,000 lives or approximately two lives every minute (1, 2) (3–5) and most of these deaths occur in low- and lower-middle-income countries.(2, 6) In India, pneumonia causes 30 million annual episodes among young children, accounting for millions of hospital admissions and 15% of under-five mortality.(3)

Reducing mortality from childhood pneumonia hinges on streamlining the care pathways.(7, 8) Prompt recognition of danger signs, appropriate management, and effective antibiotic treatment, are crucial for saving lives.(9) However, in resource-limited settings, accessing these vital interventions can be a challenge for caregivers. Financial constraints, limited knowledge about pneumonia, and long distances to healthcare facilities often deter families from seeking timely medical care.(10–15). A 2015 report by the Lancet Global Health revealed that only 54% of children exhibiting symptoms of pneumonia in low- and middle-income countries are taken outside the home for care.(16) Current coverage of treatment with effective antibiotics for children under five with pneumonia is 31% globally and 25% in India. Delayed medical attention from an appropriate provider is associated with substantial morbidity and mortality among childhood pneumonia cases. Unfortunately, studies conducted in low- and middle-income countries (LMICs) have found that healthcare workers lack expertise in initiating appropriate treatment due to insufficient training, they lack motivation due to delayed wages, and they face a shortage of medical supplies. This also results in improper management (17–19) and affects the caregivers’ and community members’ trust in health services and can affect future care-seeking behaviours.(20)

India has developed the Childhood Pneumonia Management Guidelines (CPMG) to streamline the treatment of pneumonia in children.(4) The primary aim of these guidelines is to facilitate the effective management of pneumonia symptoms such as cough, rapid breathing, and/or breathing difficulties in children under five years through community- and facility-based approaches. (4) We have previously explored barriers and facilitators to implementing the new guidelines across various tiers of the health system. Current data on barriers, behaviours, and perceptions that influence care-seeking for childhood pneumonia at the community level is currently scarce. (15, 21, 22) Such information is essential to plan effective strategies to promote health care seeking for childhood pneumonia. This study, therefore, aims to explore experiences with care-seeking for under-five children with suspected pneumonia and identify sociodemographic, and economic characteristics associated with seeking care from appropriate sources.

## METHODOLOGY

As part of a broader implementation research (IR) project, details can be found elsewhere (23), to enhance the coverage of effective pneumonia management, this study employs a mixed-methods design, integrating quantitative insights on care-seeking patterns with qualitative perspectives on underlying socio-cultural and systemic barriers, as well as experiences. The study was conducted May 1 to August 30, 2023, in Palwal district, Haryana, covering a population of 107,440 across 42 villages, which included 10 health and wellness centers, five primary health centers, four community health centers, and one district hospital.Other health care providers included private care providers, community-level health workers, chemists or pharmacies, and informal providers.[7] This study employed an Explanatory Sequential Mixed-Methods Design as part of the formative phase of a broader implementation research study. The study was conducted in two phases: the quantitative phase included a cross-sectional survey of households to identify potential under-five pneumonia cases and assess care-seeking behaviours. Following this, the qualitative phase was conducted, where in-depth interviews were carried out with purposively selected caregivers of under-five pneumonia cases. The qualitative component of this study integrated Qualitative Description (QD) and Participatory Action Research (PAR) approaches.(24, 25) The QD approach provided a clear, factual representation of caregivers’ experiences with care-seeking, capturing their perceptions and challenges without extensive theoretical interpretation.(24) Additionally, elements of PAR were embedded through active community engagement in shaping research directions, refining study tools, and informing the subsequent phases of implementation.(25) The iterative involvement of caregivers, healthcare providers, and community stakeholders, as a part of the primary implementation research, in providing feedback on pneumonia management strategies ensured that the research findings were not only descriptive but also action-oriented, contributing to policy and practice improvements in pneumonia care-seeking.(26)

For quantitative data, we obtained a list of under-five children from community health workers and conducted a cross-sectional survey using a complete enumeration technique. Relevant data were collected through house-to-house visits, covering 9,593 households with under five children. At the outset, the trained research assistants (who were university graduates by qualification) used an under-five pneumonia identification checklist (**Supplementary Annexure-I**). Children who were reported by their caregivers to have had any signs or symptoms listed in the checklist within the past 28 days were identified as potential pneumonia cases and this served as the sample for the quantitative study. For these potential under-five pneumonia cases, data were collected on household socio-demographic and economic factors along with data on possession of a “Below poverty” card, health scheme and insurance (any government health scheme or insurance e.g. Ayushman Bharat, Employees’ State Insurance scheme etc. or any private company insurance), details of pneumonia signs and symptoms, time from symptom onset to seeking care, sources of care, (**Supplementary Annexure II**) and caregivers’ previous knowledge about pneumonia (**Supplementary Annexure III**). The Below poverty card provides several benefits to households living below the poverty line. The scheme ensures that such families have free access to essential commodities, healthcare, education, housing, and employment opportunities. The information was collected using a structured questionnaire with a few open-ended questions like treatments or remedies used at home, and the symptoms caregivers believed to be indicative of pneumonia, etc. **(Supplementary Annexure-II**). This quantitative data was collected once per caregiver of under-five children, with each interview taking approximately 40–45 minutes on average. All questionnaires were translated into local language (Hindi), piloted among caregivers from April 2 to April 16, 2023, and refined based on their feedback. **(Supplementary Annexure-IV**) The quantitative data collection team was fluent in both Hindi and the local Haryanvi dialect and clarified any terms in Haryanvi during the survey, as needed, to ensure participants fully understood the questions.

The qualitative component of this study was conducted following the Standards for Reporting Qualitative Research (SRQR) guidelines. The study coordinators and investigators, who were medical doctors with specializations in public health/Ph.D. and had extensive training and experience in qualitative research methods, carried out the qualitative component using in-depth interview guides (**Supplementary Annexure V**). All guides were translated in Hindi language (**Supplementary Annexure VI**) and these investigators were proficient in Hindi language and well-versed with Haryanvi dialect. These guides focused on perceptions of pneumonia, barriers causing delays in care-seeking, and reasons for choosing specific sources of care. Purposive sampling was employed to select participants who could provide rich insights into care-seeking behaviors. Caregivers were eligible if they were fluent in the local language, Hindi or Haryanvi-dialect, to ensure effective communication. Recognizing the influence of family dynamics in decision-making, particularly in joint or three-generation households, interviews were conducted with twelve mothers, five fathers, and five grandparents. This approach aimed to capture the diverse perspectives of key family members involved in healthcare decisions, offering a more comprehensive understanding beyond the primary caregiver’s (mother’s) viewpoint. Before participation, all selected individuals were informed about the qualitative component of the study, its purpose, and their voluntary involvement. Informed consent was obtained from all participants prior to data collection. Data saturation was achieved when no new themes emerged from additional interviews, ensuring a comprehensive exploration of the research questions.

Trustworthiness of the qualitative data was ensured through multiple strategies to enhance the credibility and reliability of the findings. Investigator triangulation was employed, meaning that multiple researchers independently reviewed and analysed the data to reduce individual biases and ensure that interpretations were consistent and well-founded. This approach helped cross-verify findings and minimize any single researcher’s subjectivity influencing the conclusions. Additionally, member checking was conducted with a subset of participants, where key findings were shared with them to confirm whether the interpretations accurately represented their experiences and perspectives. This process helped ensure that the data remained true to the participants’ actual responses rather than being influenced by researchers’ assumptions. Reflexivity was also maintained throughout the research process. This involved researchers critically reflecting on their own perspectives, and potential influence on data collection and analysis. By being mindful of their preconceptions, the researchers aimed to ensure that findings were derived from the participants’ narratives rather than their own expectations. These combined measures strengthened the credibility and authenticity of the study findings.

### Outcomes

The main outcomes of the study included the determinants of healthcare-seeking in the quantitative component and caregivers’ experiences with under-five pneumonia, along with the barriers to healthcare-seeking, in the qualitative component. The outcomes included caregivers seeking care for children with pneumonia and seeking care from appropriate sources, and the time taken to seek care after the first symptoms of pneumonia appeared. The choice of source of care was assessed as a categorical outcome, grouped into appropriate (government and private health centres) and non-appropriate sources (non-RMPs, chemists etc.). Information on source of care-seeking was recorded for up to three providers: as the first source of care sought after the start of symptoms, the second source, and the third source of care. For analysis, we considered the source where caregivers sought care first as the preferred source. The operational definitions used in the analyses are detailed in **Supplementary Box 1.**

### Exposure variables

The exposure variables included sociodemographic characteristics of the under-five children and their families (such as age, gender, total under-five children in household, type of family, father’s and mother’s age, father’s and mother’s education, father’s and mother’s occupation, and household wealth index). The wealth index was developed by gathering information on household assets from the head of the household during the survey and analysing it using principal component analysis.(27) Given the small sample size, households were categorized into three equal groups: “poor”, “very poor” and “poorest” to ensure sufficient power for analysis.

### Data analysis

Quantitative data were analysed using Stata software version 16.0. Means and proportions were calculated. We utilised directed acyclic graphs (DAGs) to identify potential confounding variables for the hypothesized causal relationships with the outcome care-seeking from appropriate sources. DAG models for three exposures were constructed: Model 1 focused on wealth, Model 2 on maternal education, and Model 3 focused on maternal age as the exposure variable (**Supplementary Graph 1 to 3)**. Model I considered the type of family, mother’s age, and father’s age as confounders; Model II identified maternal age, father’s occupation, and father’s age as confounders; and in Model III considered father’s education, and father’s age were considered as confounders. Logistic regression analysis was conducted to examine the association between the exposure variables and the outcome, without and with adjustment for the confounders identified in the DAGs.

Qualitative data were managed using NVivo 1.7.1 (1534) software. Thematic analysis was performed using Braun and Clarke’s six-step framework, beginning with a deductive approach guided by the Three Delays Model, which focuses on seeking care, reaching care, and receiving care.(28, 29) This framework initially informed the coding process, allowing the researchers to systematically categorize data related to these pre-determined domains. However, as the analysis progressed, an inductive approach was also employed to identify emergent patterns and themes beyond the predefined model, capturing the nuanced realities of care-seeking behaviours and barriers in the study population. The process began with familiarization, where transcripts were thoroughly reviewed, and audio recordings were revisited to ensure an in-depth understanding of participant narratives. Systematic coding was then conducted to highlight recurring patterns related to the domains of the Three Delays Model as well as novel insights.(29, 30) These themes were iteratively reviewed and refined to ensure coherence and depth, with subthemes developed to provide further clarity.

### Ethical Approval

#### Ethical approvals

Ethical approval was granted by the ethical committees of the Society for Applied Studies (SAS/ERC/IR Pneumonia/2021), the Regional Ethics Committee of Western Norway (REK/2022/<u>531608</u>) and the World Health Organisation (WHO/ ERC.0003652). Additionally, this study has obtained approvals from the Government of Haryana state (Memo no. HSHRC/2022/505) and the health ministry steering committee (approval date: 19 Dec 2022, proposal id 2022-17596). Consent was obtained from primary caregivers and their families after reading out the information sheet to them. The participants could withdraw or request to stop recording the interviews at any time.

To enhance the transparency of our research, we have utilized the Strengthening the Reporting of Observational Studies in Epidemiology (STROBE) and Standards for Reporting Qualitative Research (SRQR) guidelines in reporting our findings.(31)

## RESULTS

After surveying 9,593 children under five, we identified 231 cases of suspected pneumonia occurring within the 28 days preceding the household visit. Out of these 231 cases, 83% (193 cases) suffered a cough, 53% (123 cases) had difficulty breathing, and 54% (125 cases) had fast breathing. Additionally, 69% (159 cases) had a temperature above 37.5°C, 25% (58 cases) suffered from vomiting, and 9% (20 cases) were unable to breastfeed. Mothers were the primary informants in 79% of cases, while both parents contributed information in 16% of the households. Two-thirds of the households possessed Below-poverty-level cards, and 35% had some form of health insurance. Around two-thirds of the pneumonia cases were male children with the average child age being 22.7 months. Educational levels varied, with 28% of mothers and 11% of fathers having less than primary education. Most mothers (97%) were housewives, while fathers primarily held private jobs (38%) or worked as daily wagers (30%). **(Table 1)**

**Table 1:**
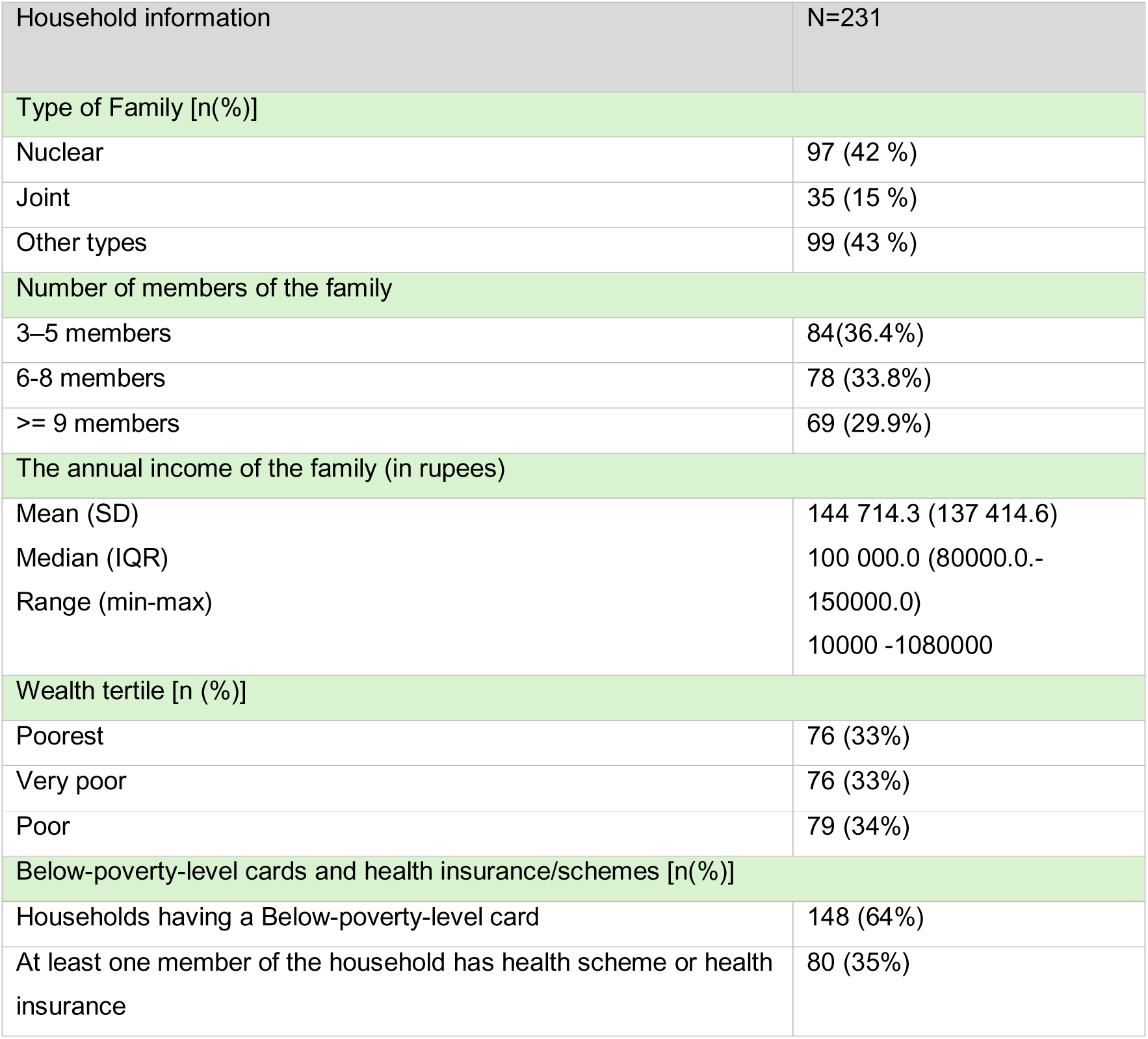
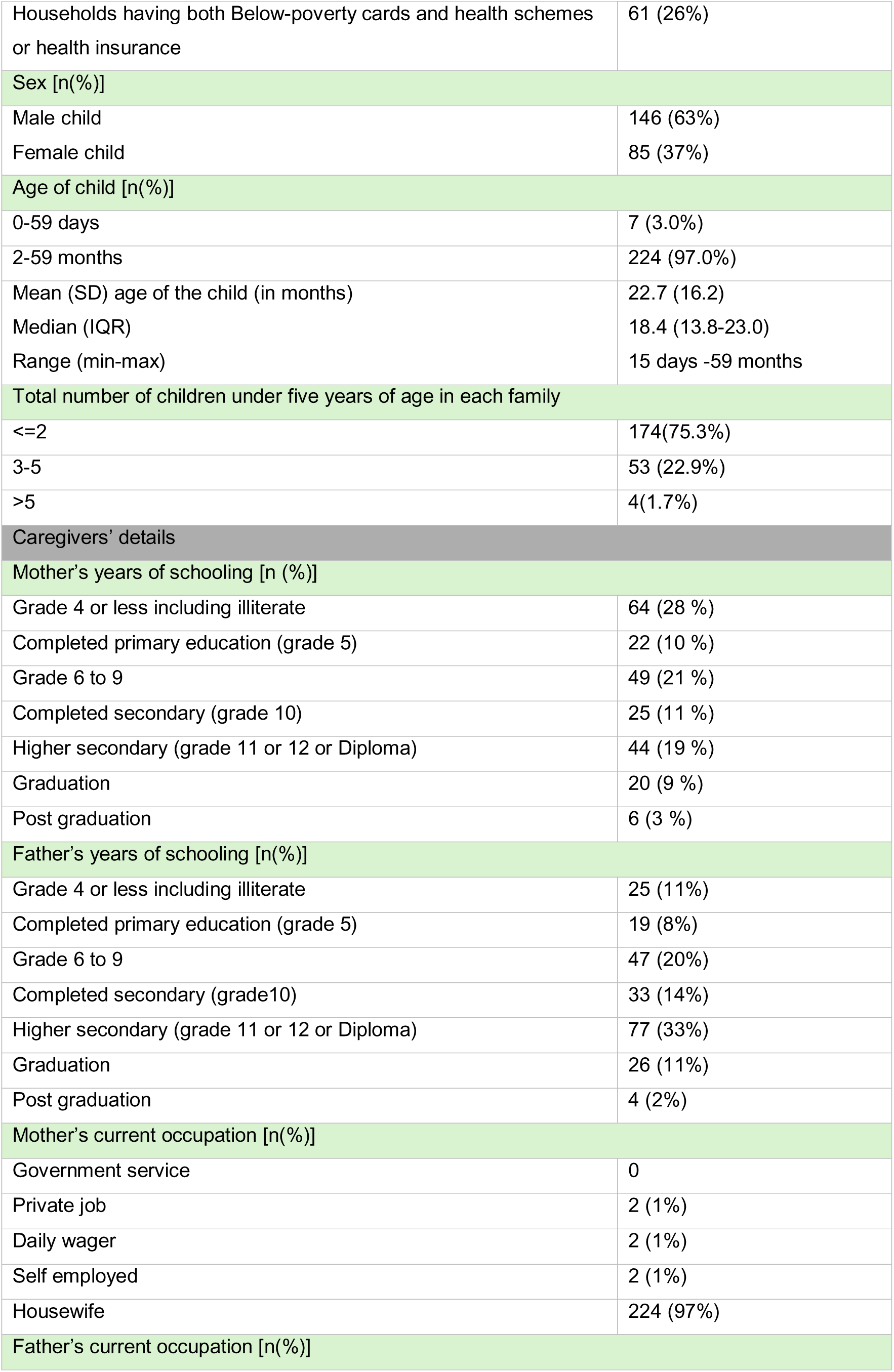
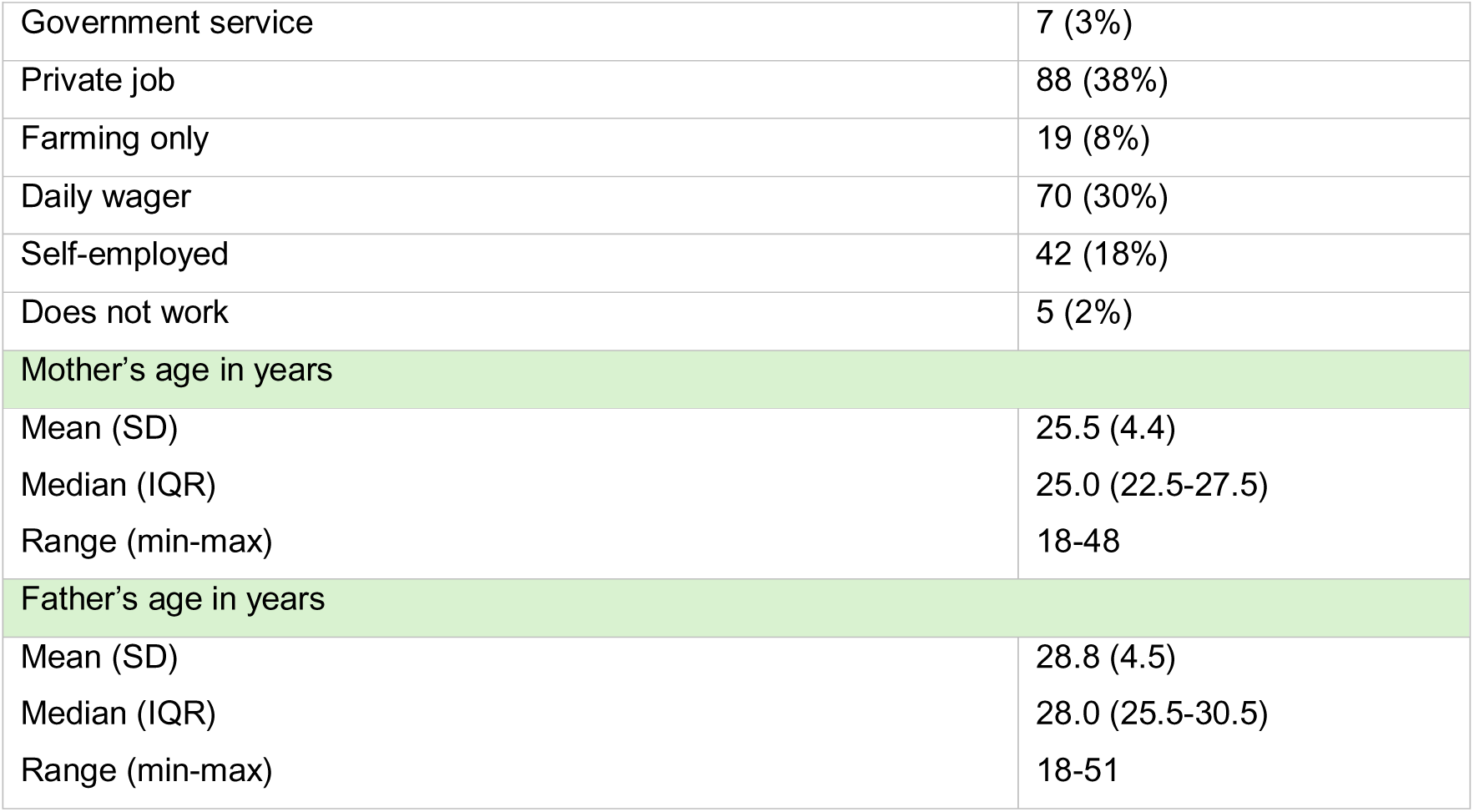
Baseline characteristics of the under-five children with pneumonia signs or symptoms in the previous 28 days.

Among the 231 under-five pneumonia cases, 97% of the caregivers sought medical care from government or private healthcare facilities or a non-RMPs. Only 3% (7 cases) did not seek any medical care. The distribution of health insurance and health care-seeking as per the availability of insurance in different socioeconomic tertiles is detailed in **Supplementary Table 1 and 2.**

Most of the caregivers sought care from non-registered practitioners, and one-fourth utilized private facilities, which included private hospitals/nursing homes. Government healthcare options were less frequently used and 4 out of 224 caregivers i.e., around 2%, directly sought assistance from chemists for obtaining medicines. (**Table 2)**

**Table 2:** First source of care sought among caregivers of under-five children with suspected pneumonia.

| Sources of care | N=224<br>n (%) |
| --- | --- |
| Government | 8 (3.6%) |
| Accredited social health activist | 1 (0.4%) |
| Auxiliary nurse midwives/community health officers (at health and wellness centers) | 2 (0.9%) |
| Medical officers (at primary or community health centers) | 2 (0.9%) |
| District hospital | 3 (1.3%) |
| Private | 57 (25.4%) |
| Private hospitals/Nursing homes | 39 (17.4%) |
| Private Clinics (In-charge with an MBBS/MD degree) | 13 (5.8%) |
| Alternative Medicine<br>(In-charge with a BAMS and BHMS degree) | 5 (2.2%) |
| Non-registered medical practitioner | 159 (71%) |
| Private (Degree of provider unknown) | 155 (69.2%) |
| Chemist | 4 (1.8%) |

MBBS: Bachelor of Medicine and Bachelor of Surgery; MD: Doctor of Medicine; BAMS: Bachelor of Ayurvedic Medicine and Surgery; BHMS: Bachelor of Homeopathic Medicine and Surgery.

The duration within which caregivers sought care for under-five children with suspected pneumonia is detailed in **Table 3**. Out of the children with suspected pneumonia, children in the poorest families were most likely to receive no treatment (**Supplementary Graph 4**). Across all economic groups, families most often turned to non-RMPs for pneumonia treatment. This tendency was particularly pronounced among the poorest families. The poor families were more likely to use private healthcare than the poorest and very poor families, and the government facilities were the least preferred facilities for care in all the wealth categories.

**Table 3:** Duration within which caregivers sought care for under-five children with suspected pneumonia (N=224)

| Care-seeking from appropriate sources (n=65) |  |  |  |
| --- | --- | --- | --- |
| Duration | N (n%) | Mean (SD) age of child in months | Median (IQR) age of child in months |
| Within 24 hours | 51 (78.4) | 26.4 (17.2) | 20.1(2.9 to 58.5) |
| >24 and <= 48 hours | 12 (18.4) | 22.7(17.1) | 14.2(2.7 to 48.8) |
| >48 hours | 2 (3.1) | 12.2 (2.5) | 12.2 (10.9-13.4) |
| Care-seeking from non-appropriate* sources (n=159) |  |  |  |
| Duration | N (n%) | Mean (SD) age of child in months | Median (IQR) age of child in months |
| Within 24 hours | 136 (85.5) | 26.2 (15.7) | 24.6(4.2 to 59.3) |
| >24 and <= 48 hours | 16 (10.1) | 24.43 (17.6) | 20.4 (7.9 to 56.9) |
| >48 hours | 7 (4.4) | 26.4(16.3) | 16.3 (7.7 to 57.6) |

In the multivariable model, higher maternal education (diploma level or above) was associated with an increased likelihood of seeking appropriate care for under-five pneumonia (**Table 4)**. The least poor group were also more likely to seek care from appropriate sources compared to the poorest group. Maternal age was inversely associated with appropriate care-seeking, but the wide confidence intervals suggest that these results were uncertain. (Table 4) An overview of the qualitative findings is provided in **Table 5**. Caregivers often misinterpreted symptoms such as difficulty in breathing or chest in-drawings as signs of common cold or seasonal ailments, resulting in delays in seeking timely medical care. Some families, particularly mothers, perceived these symptoms as indicative of a disease but believed they were unlikely to lead to serious illness, which contributed to delays in care-seeking. More concerningly, certain families, especially those with grandparents as household heads, viewed these signs and symptoms as natural adaptations of the child’s body to seasonal changes, such as the transition from autumn to winter, and did not recognize them as indicators of illness. Grandmothers often held strong beliefs that such symptoms, like those experienced by their own children in the past, could be adequately managed with home remedies, negating the need for professional medical attention. Furthermore, findings from a subset of households indicated that poor household environmental conditions, such as exposure to indoor smoke, exacerbated respiratory symptoms, and the team observed that there was limited awareness of strategies to mitigate these risks.

**Table 4:** Associations between mothers’ sociodemographic characteristics and care-seeking from appropriate sources for under-five pneumonia.

| Variables |  | Care-seeking from appropriate sources |  |  |
| --- | --- | --- | --- | --- |
|  | n (%) | Unadjusted<br>odds ratio<br>[95% C.I.] | Adjusted<br>odds ratio<br>[95% C.I.) | Variables<br>adjusted for |
| Wealth tertiles |  |  |  |  |
| Poorest | 10/76 (13.2) | Reference |  | Mother's age,<br>Father's age<br>Type of family |
| Very Poor | 21/76 (27.6) | 0.9<br>[0.1,1.8] | 1.0<br>[0.1,1.9] |  |
| Poor | 34/79 (43.1) | 1.6<br>[0.8,2.4] | 1.7<br>[1.0,2.6] |  |
| Mother's education (in years) |  |  |  |  |
| Primary education or illiterate | 10/87 (11.5) | Reference |  | Maternal age,<br>father's<br>occupation,<br>and father's<br>age |
| Above primary to secondary education | 22/74 (29.7) | 3.3<br>[1.4,7.4] | 3.1<br>[1.3,7.3] |  |
| Diploma to above graduation | 33/70 (47.1) | 6.7<br>[3.1,10.1] | 6.5<br>[2.7,9.7] |  |
| Mother's age (in years) |  |  |  |  |
| Less than or equal to 21 years | 10/27 (37.0) | Reference |  | Father's<br>education and<br>father's age. |
| 22 to 25 years | 35/110 (31.2) | 0.8<br>[0.3,1.9] | 0.9<br>[0.3,2.5] |  |
| 26 to 30 years | 17/76 (22.4) | 0.5 | 0.7 |  |
|  |  | [0.2,1.3] | [0.2,2.3] |  |
| 31 years and above | 3/18 (16.7) | 0.3<br>[0.1,1.5] | 0.6<br>[0.1,3.6] |  |

**Table 5:**
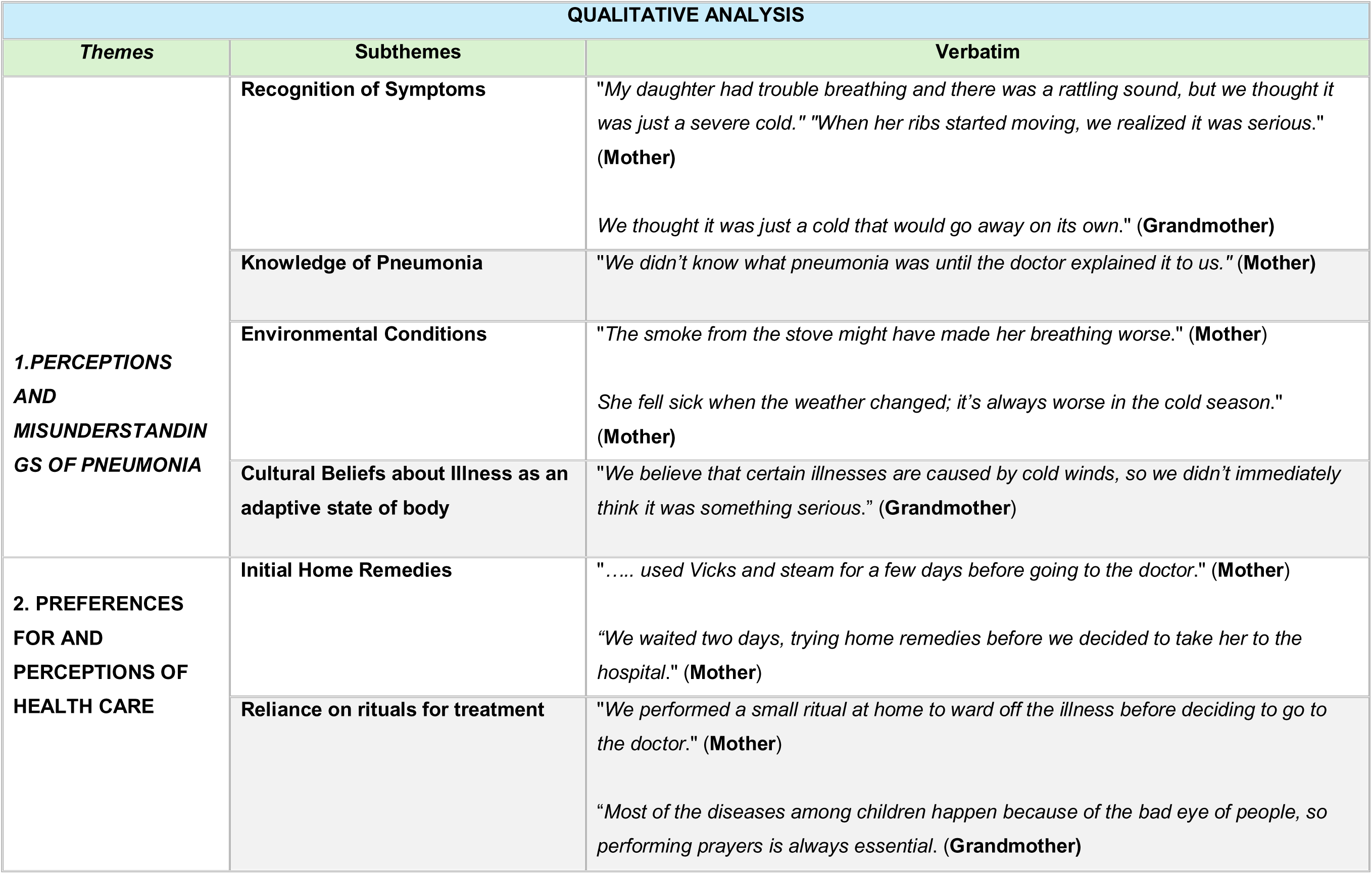
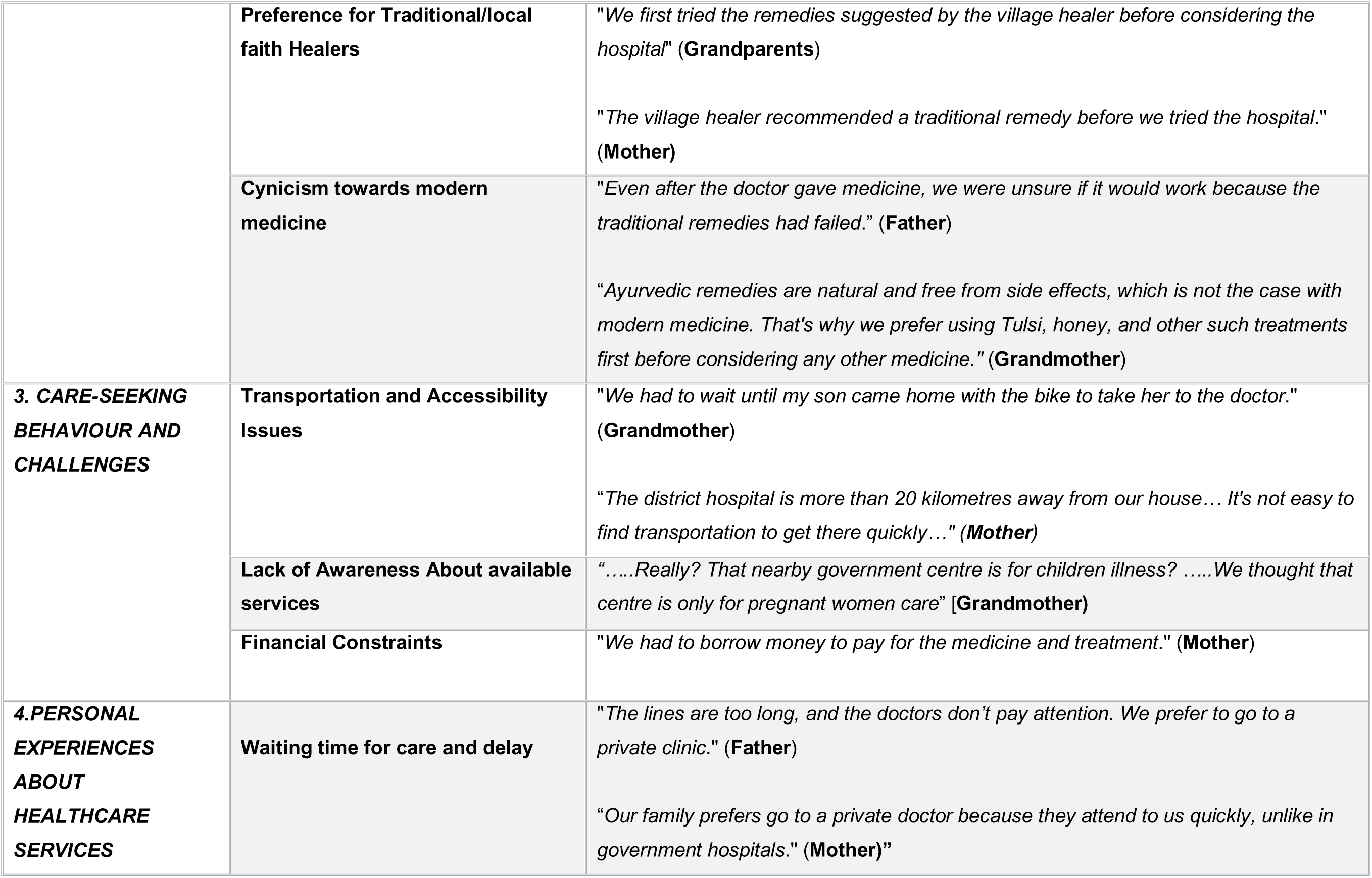
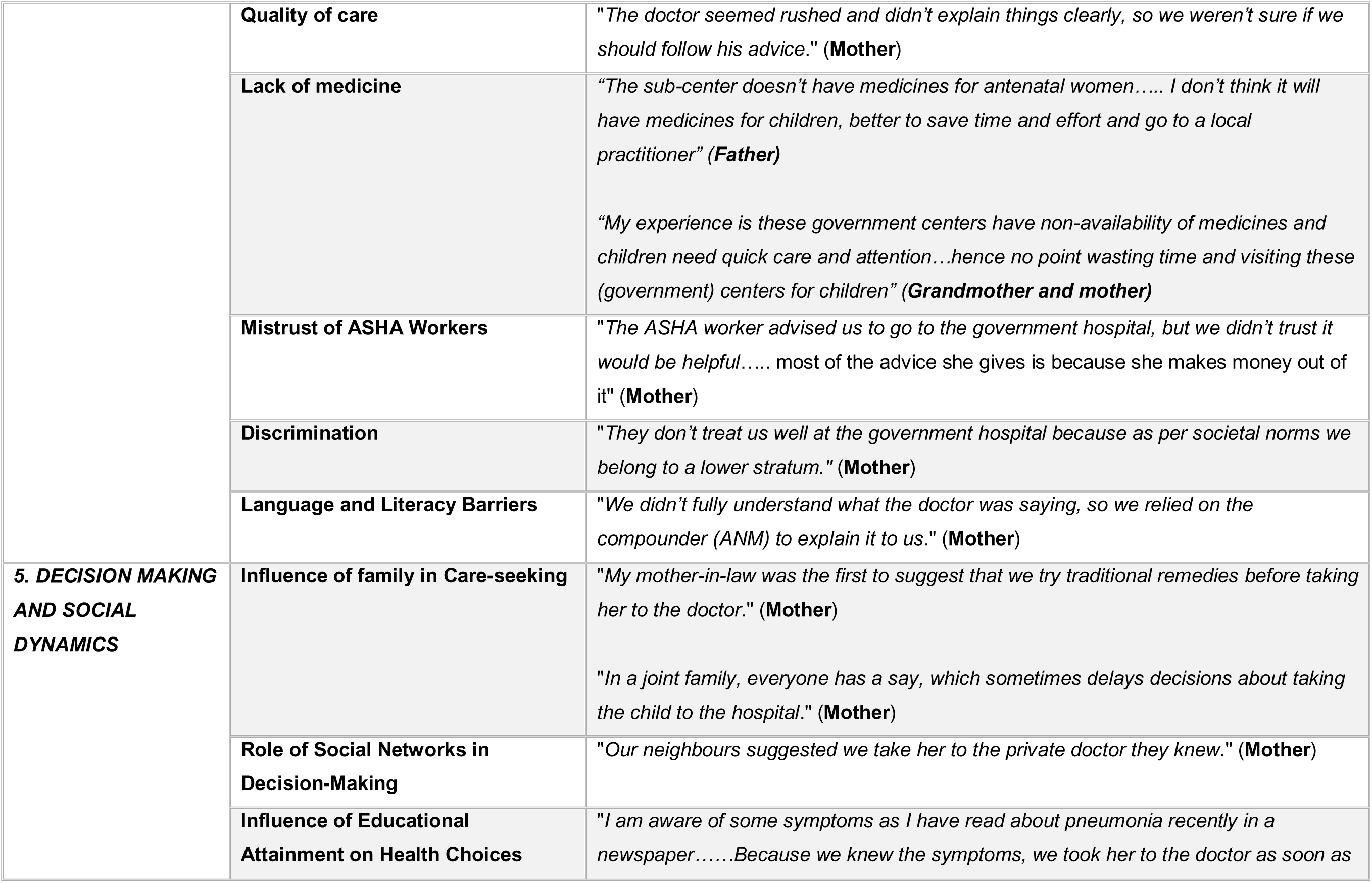
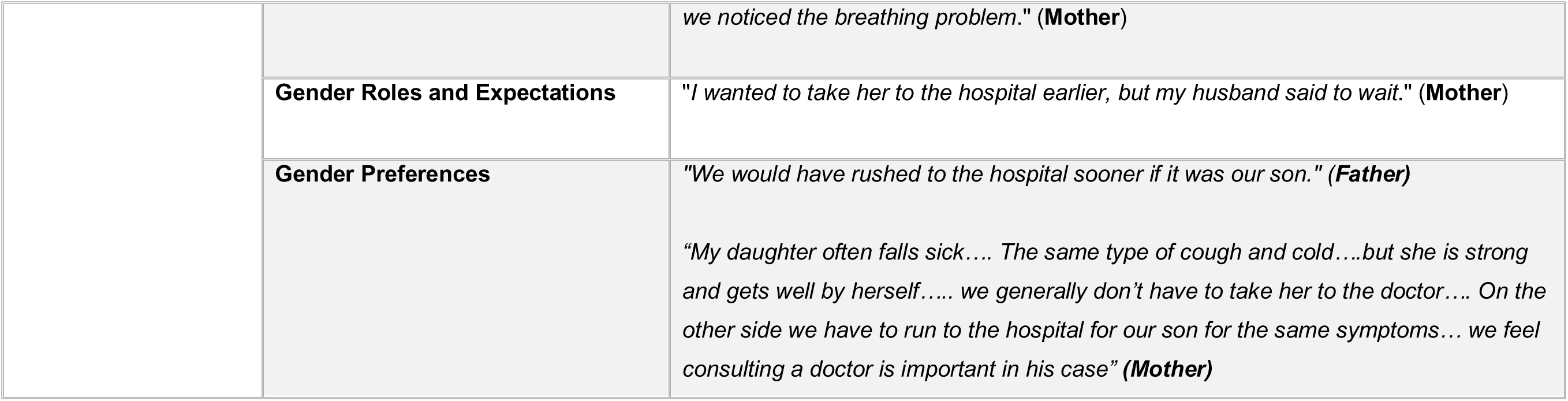
Thematic Analysis of Care-Seeking Behaviour for Childhood Pneumonia in Rural India.

Caregivers demonstrated a strong reliance on home remedies and traditional healers, often delaying formal healthcare interventions until traditional methods had failed. Among many households, initial treatment involved rituals and superstitions, stemming from a belief that illnesses were caused by the “evil eye” or negative influences from others in the community. This practice was widespread. Additionally, significant cynicism toward modern medicine was observed in some families. This scepticism stemmed from a belief that allopathic treatments were inherently associated with side effects, leading to hesitation in fully trusting modern medical care. Government healthcare facilities were often viewed as inefficient due to long waiting times and poor service quality. The perceived unavailability of medicines further deterred families, who believed essential medicines for children would also be lacking. While ASHA workers disseminated information about childhood illnesses and appropriate care options, mistrust in their advice undermined community confidence in public health services. Additionally, sporadic reports of societal-class-based discrimination discouraged families from seeking care at government facilities. These factors collectively led families to favor private providers, despite the higher financial burden and availability of government healthcare services. Seeking care from private practitioners for childhood illnesses could often cost a family approximately six to seven dollars per visit per child. The perceived efficiency, responsiveness, and individualized attention offered by private practitioners were strong factors driving their preference.

Logistical barriers, such as limited transportation options and financial constraints, were significant factors delaying timely care. Many caregivers reported being unaware of government health centers providing paediatric care, assuming these facilities catered only to antenatal and postnatal services.

Family dynamics significantly influenced care-seeking decisions, with grandparents often advocating for traditional remedies, contributing to delays in seeking appropriate medical care. In joint family structures, the involvement of multiple members in decision-making further slowed the process of determining when and where to seek care. Social networks, including neighbours and community members, also shaped healthcare choices within households. Traditional gender roles played a pivotal role, as women frequently deferred healthcare decisions to male family members. Additionally, caregivers with higher educational attainment exhibited better health literacy, enabling them to recognize symptoms promptly and seek timely medical intervention. Gender biases also played a role in shaping care-seeking behaviour, with boys often receiving quicker medical attention compared to girls. Families perceived boys as being more vulnerable to illness and prioritized their health needs over those of girls. In contrast, girls were often considered to be inherently stronger and more resilient, leading to delayed care-seeking for female children.

## Discussion

Of the 231 suspected cases of under-five pneumonia, 97% of caregivers sought medical care, but the majority relied on Non-RMPs, with only a minority accessing appropriate sources.

Socioeconomic factors, such as higher maternal education and being in the least poor wealth group, were associated with a greater likelihood of seeking care from appropriate sources. The qualitative findings revealed that delays in treatment were often due to a perception that pneumonia is not a life-threatening disease, reliance on home remedies, consultations with traditional healers, and financial constraints. Additional barriers included mistrust in government healthcare services regarding quality of services provided, social dynamics complicating decision-making, and, in some cases, gender biases that delayed care-seeking.

Logistical barriers and dependence on male family members for transportation, significantly delayed care, as highlighted in previous studies.(32) In this community, women making healthcare decisions independently or taking their children to healthcare practitioners without male or any other family-member accompaniment are often perceived as culturally inappropriate behaviour. To avoid social repercussions and maintain their own reputation, women typically conform to established socio-cultural norms, which can sometimes hinder timely care-seeking. Additionally, most women in this community were found to be less educated and unemployed, which may have contributed to their lack of confidence and hesitation in independently approaching the tertiary level or distant healthcare centers.(33) The Ayushman Bharat program in India aims to address such challenges by establishing Health and Wellness Centres (HWCs) for decentralized healthcare.(34) However, many caregivers reported that these centers were predominantly utilized for antenatal care services. Caregivers expressed willingness to use HWCs for under-five care if such services were made available. Raising awareness about the services offered at HWCs and ensuring their functionality remain critical for enhancing access, especially in rural areas where transportation challenges persist. In addition, empowering women to make independent healthcare decisions could help.

Financial constraints emerged as a significant barrier, with many families delaying treatment until they could borrow money. This burden was particularly acute for those relying on private practitioners and hospitals due to perceived gaps in government hospitals such as the frequent unavailability of essential medicines and the lack of diagnostic facilities. Although healthcare services and drugs for children are intended to be free in government facilities, these shortcomings force families to incur out-of-pocket expenses. Similar patterns have been reported in other LMICs settings where financial limitations significantly delayed care-seeking for childhood illnesses.(7, 8, 35, 36) Financial restrictions may also lead to prioritizing treatment for male children, as observed in this study. While boys were often perceived as more vulnerable than girls, this perception may reflect deeper sociocultural norms. In many Indian settings, boys are traditionally regarded as more valuable to the family due to their perceived role in ensuring future welfare, performing death rites, and upholding family prestige. Studies from India have documented that families often prioritize healthcare and financial expenditures for male children over females, leading to delayed or inadequate care for girls.(37, 38) Such findings have been reported in studies conducted in other LMICs too.(32, 39) The preference for home remedies as the initial choice of care and reliance on spiritual healers probably reflected deeply rooted cultural norms and spiritual explanations for illness but may also be partially attributed to financial constraints. With an average monthly household income of around 158 dollars and limited awareness of the availability of free medicines and consultations at nearby government facilities, families are naturally inclined to choose cost-effective options. Consequently, financial limitations indirectly shape and restrict their primary healthcare choices. Targeted interventions to address financial barriers, by expanding the reach and awareness of government healthcare programs, such as Ayushman Bharat, to ensure that essential medicines and diagnostic facilities are consistently available in government facilities, is critical. (34) Also, increasing awareness on free transportation facilities to hospital for sick under-five children available under national ambulance policy, could alleviate the economic burden on families.(40) Strengthening financial literacy and providing clear information about available health insurance options can further mitigate financial constraints. Additionally, introducing community-based health financing schemes or microloans to support families during medical emergencies could reduce delays in seeking care. These measures could not only improve equitable access to care for both boys and girls but also reduce the delays in seeking timely healthcare from appropriate healthcare facilities across socio-economic groups.

There was a clear socioeconomic gradient in health insurance coverage, but our findings also indicated that health insurance coverage did not eliminate all financial barriers. Several factors may contribute to more than half of the insured caregivers opting for care from non-RMPs, like perceived complexity of insurance schemes, indirect costs like transportation and lost wages, and mistrust in government facilities, discouraging families from using insured services.(41, 42).

Another critical issue identified was the nature of counselling provided at government healthcare facilities. Counselling sessions were often one-sided and lacked meaningful interaction. Healthcare providers of the government centres frequently failed to dedicate sufficient time to understand the perspectives and concerns of beneficiaries, resulting in suboptimal quality of care. Lack of engagement and personalized attention further dissuades beneficiaries from utilizing public health services and drive families toward private providers despite higher costs, as also seen in studies from Peru.(36) The preference for non-RMPs may stem from their accessibility, convenience, and established community relationships. Enhancing the quality and reliability of government services is crucial to rebuilding trust and encouraging utilization. However, the qualitative findings revealed that mistrust in government services also often arose from community attitudes and rumours rather than direct personal experiences. The erosion of trust in ASHAs is partly driven by the perception that they prioritize national health programs that offer financial incentives over those that do not. Since many ASHAs focus more on activities linked to incentives, community members may feel that their engagement is transactional rather than driven by a commitment to overall health promotion. This perception can undermine the credibility of ASHAs and affect participation in non-incentivized health initiatives.Additionally, rumours casting doubt on their commitment to community welfare, could have long-term detrimental effects on the healthcare system. Efforts to improve public healthcare delivery, build community trust, e.g. through clearly demonstrating efforts to be responsive and accountable, and ensuring the consistent availability of essential medications can make formal healthcare more accessible and appealing to the most vulnerable households. This may be combined with addressing gender biases through community awareness programs, such as engaging with frontline health workers like ASHAs in counselling about girl children’s health, to achieve more equitable health outcomes for all children.

The perception that pneumonia is not life-threatening, as observed in this study, significantly contributes to delays in seeking formal care. Similar findings from other resource-limited settings, where misbeliefs about illness severity led to delayed healthcare-seeking have been found.(7, 8, 43) The interplay of cultural beliefs, mistrust of modern medicine, and perceptions of healthcare quality highlights the complex factors shaping care-seeking behaviour in these communities. Addressing these challenges requires educational campaigns emphasizing the severity of pneumonia and the risks of delayed care. Such interventions should target entire households, recognizing the influential role of family members, particularly grandparents, in healthcare decision-making.

## Strengths and Limitations

This study’s mixed-methods design is a major strength, providing an understanding of many aspects of care-seeking behaviour. The inclusion of multiple family members, including mothers, fathers, and grandparents, offered diverse perspectives that enhanced our understanding of healthcare decision-making. Additionally, this study, as part of the formative research within a broader implementation research (IR), integrates structured community engagement in shaping implementation strategies. The findings from this phase will inform the development of the initial implementation model, while the broader IR will facilitate the ongoing involvement of caregivers, family members, community representatives, and healthcare providers in refining and adapting strategies using structured feedback mechanisms (**Supplementary Annexure-III and VII).** Additionally, the findings will support in preparing a logic model (44) developed using the implementation research principles and tools.(45, 46) This strong engagement of beneficiaries, alongside active participation from community representatives and healthcare providers, enhances the study’s relevance, sustainability, and future impact by ensuring community-driven, contextually appropriate strategies for improving pneumonia care-seeking behaviours and effective management in rural India.

The reliance on self-reported data introduced the possibility of recall bias, especially concerning the timing and source of care-seeking. Additionally, symptoms may have been over-reported or inaccurately recalled, inflating the number of suspected pneumonia cases. This issue is compounded by the low specificity of the pneumonia definition used, which includes a broad range of symptoms without using investigations like chest X-rays or laboratory tests. Consequently, it might capture many cases of other respiratory illnesses, leading to an overestimation of pneumonia cases. Furthermore, the qualitative component relied on purposive sampling, which enabled an in-depth exploration of diverse perspectives. However, this approach may not have fully captured the experiences of less vocal caregivers or those facing local language barriers, who might encounter different challenges in care-seeking. Social desirability bias may have also influenced responses, as some caregivers could have provided answers they perceived as favourable rather than their actual experiences. Additionally, while this study primarily focused on community-level care-seeking experiences, incorporating data from healthcare providers and health system stakeholders could have further enhanced validity and facilitated triangulation of findings. However, this was beyond the scope of this manuscript. To address this gap, a separate manuscript exploring health system challenges and healthcare provider perspectives on pneumonia management has been published.(26) The findings of this study are also specific to the Palwal district, limiting their generalizability to regions with different healthcare infrastructures or cultural contexts.

## Conclusion

This study highlights the complex socio-economic, cultural, and systemic factors that shape care-seeking behaviour for childhood pneumonia in rural North India. Delays in seeking appropriate care are driven by poverty, reliance on non-RMPs, social dynamics, in-adequate awareness and knowledge of pneumonia, gender biases and mistrust in public healthcare systems. These findings contribute to the growing body of evidence on care-seeking behaviour in LMICs and underscore the need for multifaceted approaches to improving healthcare access.

## Declarations

## Consent for publication

Not applicable.

## Conflict of interests

The authors declare that they have no competing interests.

## Funding

The study was funded by the Bill & Melinda Gates Foundation (#INV-008068) through a grant to the World Health Organization. The funders had no role in the study design or in the collection, analysis, or interpretation of the data. The funders did not write the report and had no role in the decision to submit the paper for publication.

## Disclaimer

The authors alone are responsible for the views expressed in this article and they do not necessarily represent the views, decisions, or policies of the institutions with which they are affiliated.

## Data availability statement

The data used and/or analysed during the current study are available from the corresponding author/s Barsha Gadapani Pathak and/or and Sarmila Mazumder and on reasonable request.

## Acknowledgements

The authors would like to express sincere gratitude to all the under-five caregivers who participated in this study and shared their experiences. They would also like to thank Mr. Hashmi and Dr. Manish Kumar for coordinating research activities and Ms Arti, Ms Kavita, and Ms Dipti for assisting in the data generation process.

## Ethical approvals

Ethical approval was granted by the ethical committees of the Society for Applied Studies (**SAS/ERC/IR Pneumonia/2021**), the Regional Ethics Committee of Western Norway (**REK**/**2022/<u>531608</u>**) and the World Health Organisation (**WHO/ ERC.0003652**). Additionally, this study has obtained approvals from the Government of Haryana state (**Memo no. HSHRC/2022/505**) and the health ministry steering committee (**approval date: 19 Dec 2022, proposal id 2022-17596)**. Consent was obtained from primary caregivers and their families after reading out the information sheet to them. The participants could withdraw or request to stop recording the interviews at any time.

## Notes

### Competing Interest Statement

The authors have declared no competing interest.

### Author Declarations

Ethical approval was granted by the ethical committees of the Society for Applied Studies (SAS/ERC/IR Pneumonia/2021), the Regional Ethics Committee of Western Norway (REK/2022/531608) and the World Health Organisation (WHO/ ERC.0003652). Additionally, this study has obtained approvals from the Government of Haryana state (Memo no. HSHRC/2022/505) and the health ministry steering committee (approval date: 19 Dec 2022, proposal id 2022-17596). Consent was obtained from primary caregivers and their families after reading out the information sheet to them. The participants could withdraw or request to stop recording the interviews at any time.

